# Repurposing Non-pharmacological Interventions for Alzheimer’s Diseases through Link Prediction on Biomedical Literature

**DOI:** 10.1101/2023.05.15.23290002

**Authors:** Yongkang Xiao, Yu Hou, Huixue Zhou, Gayo Diallo, Marcelo Fiszman, Julian Wolfson, Halil Kilicoglu, You Chen, Chang Su, Hua Xu, William G. Mantyh, Rui Zhang

## Abstract

Recently, computational drug repurposing has emerged as a promising method for identifying new pharmaceutical interventions (PI) for Alzheimer’s Disease (AD). Non-pharmaceutical interventions (NPI), such as Vitamin E and Music therapy, have great potential to improve cognitive function and slow the progression of AD, but have largely been unexplored. This study predicts novel NPIs for AD through link prediction on our developed biomedical knowledge graph. We constructed a comprehensive knowledge graph containing AD concepts and various potential interventions, called ADInt, by integrating a dietary supplement domain knowledge graph, SuppKG, with semantic relations from SemMedDB database. Four knowledge graph embedding models (TransE, RotatE, DistMult and ComplEX) and two graph convolutional network models (R-GCN and CompGCN) were compared to learn the representation of ADInt. R-GCN outperformed other models by evaluating on the time slice test set and the clinical trial test set and was used to generate the score tables of the link prediction task. Discovery patterns were applied to generate mechanism pathways for high scoring triples. Our ADInt had 162,213 nodes and 1,017,319 edges. The graph convolutional network model, R-GCN, performed best in both the Time Slicing test set (MR = 7.099, MRR = 0.5007, Hits@1 = 0.4112, Hits@3 = 0.5058, Hits@10 = 0.6804) and the Clinical Trials test set (MR = 1.731, MRR = 0.8582, Hits@1 = 0.7906, Hits@3 = 0.9033, Hits@10 = 0.9848). Among high scoring triples in the link prediction results, we found the plausible mechanism pathways of (Photodynamic therapy, PREVENTS, Alzheimer’s Disease) and (Choerospondias axillaris, PREVENTS, Alzheimer’s Disease) by discovery patterns and discussed them further. In conclusion, we presented a novel methodology to extend an existing knowledge graph and discover NPIs (dietary supplements (DS) and complementary and integrative health (CIH)) for AD. We used discovery patterns to find mechanisms for predicted triples to solve the poor interpretability of artificial neural networks. Our method can potentially be applied to other clinical problems, such as discovering drug adverse reactions and drug-drug interactions.

## Introduction

Alzheimer’s disease (AD) and related dementias (ADRD) are chronic and multifactorial neurodegenerative disorders that affect cognition, behavior, functional ability and memory of affected individuals ^1^. As of 2020, the worldwide prevalence of ADRD was approximately 50 million, and this number is expected to increase to 152 million by 2050, representing a significant and growing public health challenge ^2^. The high prevalence of ADRD has significant economic, medical, and social consequences for society. In 2019, the global economic burden of ADRD was estimated to be $2.8 trillion, and this burden is projected to increase to $16.9 trillion by 2050 ^3^. Despite significant advances in our understanding of the etiology and drug targets of AD/ADRD, effective prevention and treatment of these conditions remain elusive. Several medications, including lecanemab ^4^ and aducanumab ^5^, have been developed based on well-defined concepts and hypotheses about the etiology and drug targets of AD/ADRD. These medications are thought to reduce the pathological progression of the disease; however, their treatment effect is limited ^6^. This suggests that our understanding of the pathogenesis of Alzheimer’s disease is incomplete, and that novel unbiased approaches are needed to discover new therapies.

AD is a complex and multifactorial disorder that poses significant challenges to drug discovery research. Despite significant progress in this field, there remains an unmet need for effective treatments, prevention, or interventions to slow down the progression of AD ^7^. Pharmacological interventions (PI) have demonstrated improvements in cognitive function, albeit with adverse side effects such as nausea, weight loss, leg cramps, and increased mortality risk ^8,9^. On the other hand, non-pharmacological interventions (NPI) including sleep ^10,11^, diet ^12^, dietary supplements ^13^, aerobic exercise ^14^, aromatherapy ^15^, light therapy ^16^ and cognitive training ^17^ are widely used by healthcare consumers to enhance their well-being and manage diseases. Thus, NPIs represent a promising, versatile, and potentially cost-effective approach to improve outcomes and quality of life for patients with dementia ^18^. Recent studies have demonstrated that certain NPIs may be protective against cognitive decline in individuals with positive biomarkers and cognitive impairment ^19^. For example, aerobic exercise has been shown to benefit various aspects of cognition, including the stabilization of Mini-Mental State Examination (MMSE) scores, as well as improvements in attention, memory, and recognition ^20,21^. Cognitive decline may also be attenuated by factors such as improved nutrition, appropriate dietary supplements, mental exercise, and social activities ^22^. Notably, multimodal NPIs have shown promise in improving cognitive function ^23,24^. However, a comprehensive understanding of the effects of NPI, as well as the potential synergistic effects of PI and NPI for AD/ADRD, remains lacking.

In recent years, the analysis of existing data on drugs and diseases has emerged as a promising approach for discovering new therapeutic potentials of existing drugs and identifying treatments for refractory diseases, a practice commonly referred to as drug repurposing ^25^. Text mining is a popular data mining approach for drug repurposing due to the rapidly increasing volume of biomedical and pharmaceutical research literature. A vast number of semantic relations between biomedical entities can now be extracted from this literature. Knowledge graphs (KGs), which are heterogeneous networks, can be utilized to store, manage and represent these semantic relations. KGs can be tools used to model entities and their relationships, and the network structure of KGs can be leveraged to generate hypotheses by utilizing graph theory concepts and methods ^25^. In biomedical knowledge graphs (BKGs), nodes signify biomedical entities, and edges represent the relationships between two entities ^26^. BKGs can provide solutions to practical problems in the biomedical domain. For instance, the SuppKG, a Dietary Supplement domain knowledge graph, can identify interactions between drugs and dietary supplements through discovery patterns ^27^. Link Prediction (LP) for knowledge graphs (also known as knowledge graph completion) is the task of inferring missing or potential relations between entities in a knowledge graph ^28^.

The LP for Semantic MEDLINE Database (SemMedDB) ^29^ has been found to be effective for drug repurposing for COVID-19 ^30^. To address the current lack of research exploring novel NPIs for AD, we first trained and evaluated various LP strategies (e.g., embedding-based, neural network based models). The best-performing model was further utilized to predict NPIs that may have the potential to prevent AD. The NPIs include natural products (e.g., dietary supplements (DS)) and complementary and integrative health (CIH), which are identified using self-contained information in SuppKG and a CIHLex we previously created ^31^, respectively. Subsequently, discovery patterns ^32^ are employed to generate mechanism pathways for candidates with high scores (i.e., high likelihood), and these pathways are evaluated by domain experts. Our contribution includes creating NPI resources and developing an innovative framework to predict NPIs that may potentially be repurposed for AD. To our best of knowledge, this is the first study to discover NPIs for AD. The developed framework can be applied to NPI discovery for other diseases.

## Results

### ADInt Statistics

The comprehensive AD Intervention knowledge graph (called ADInt) encompasses 162,213 entities across 113 UMLS semantic types, which after further identification include 25,604 Drugs, 16,474 Diseases, 46,060 Genes and Proteins, 2,525 DS, and 128 CIH. Furthermore, ADInt comprises 1,017,319 triples, capturing 15 distinct relation types such as INTERACTS_WITH, AFFECTS and TREATS. Detailed statistics can be found in Table 1.

**Table 1:** The frequency and the proportion of relation types in ADInt.

| Relations | Counts (%) | Relations | Counts (%) |
| --- | --- | --- | --- |
| COEXISTS_WITH | 332,441 (32.68) | DISRUPTS | 23239 (2.28) |
| INTERACTS_WITH | 209,458 (20.59) | AUGMENTS | 21,912 (2.15) |
| AFFECTS | 96,804 (9.52) | PRODUCES | 21,828 (2.14) |
| TREATS | 90,812 (8.93) | PREDISPOSES | 13,509 (1.33) |
| CAUSES | 76,236 (7.49) | PREVENTS | 12,258 (1.20) |
| ASSOCIATED_WITH | 46,126 (4.53) | COMPLICATES | 3,519 (0.35) |
| INHIBITS | 39,158 (3.85) | MANIFESTATION_OF | 1,926 (0.19) |
| STIMULATES | 28,093 (2.76) |  |  |
| **TOTAL** |  | 1,017,319 |  |

### Performance of LP models

Table 2 presents the performance obtained by various LP methods using the metrics Mean Rank (MR), Mean Reciprocal Rank (MRR), and Hits@k (k = 1, 3, and 10) ^33^. A well-performing model should exhibit a low MR score and high MRR and Hit@k scores. The results demonstrate that the R-GCN model outperforms the other models in all metrics, followed by the TransE model. Notably, the CompGCN model performs the lowest performance across all metrics.

**Table 2:** The metrics of link prediction results for different models on integrated knowledge graph, ADInt, by time slicing evaluation.

|  | TransE | RotatE | DistMult | Complex | RGCN | CompGCN |
| --- | --- | --- | --- | --- | --- | --- |
| Hits@1 | 0.1770 | 0.1786 | 0.1109 | 0.1062 | **0.4112** | 0.0906 |
| Hits@3 | 0.3242 | 0.3055 | 0.2586 | 0.2467 | **0.5058** | 0.1509 |
| Hits@10 | 0.5996 | 0.5340 | 0.5921 | 0.5854 | **0.6804** | 0.4267 |
| MRR | 0.3109 | 0.2987 | 0.2547 | 0.2479 | **0.5007** | 0.1973 |
| MR | 8.861 | 10.11 | 9.278 | 9.380 | **7.099** | 10.26 |

Additionally, Table 3 reports evaluation results of the trained models on the Clinical Trials dataset. The findings show that the R-GCN model performs the best across all metrics. In this case, some metrics of the RotatE model (Hits@3 = 0.6320, Hits@10=0.8107, MR=5.228) are better than TransE (Hits@3 = 0.6294, Hits@10=0.7621, MR=5.417). Collectively, from both evaluation results presented in Table 2 and Table 3, the R-GCN model exhibits the best performance, with the lowest MR and the highest MRR, Hits@1, Hits@3, and Hits@10 among the considered models. Thus, we used the R-GCN for further knowledge discovery of NPIs on AD prevention.

**Table 3:**
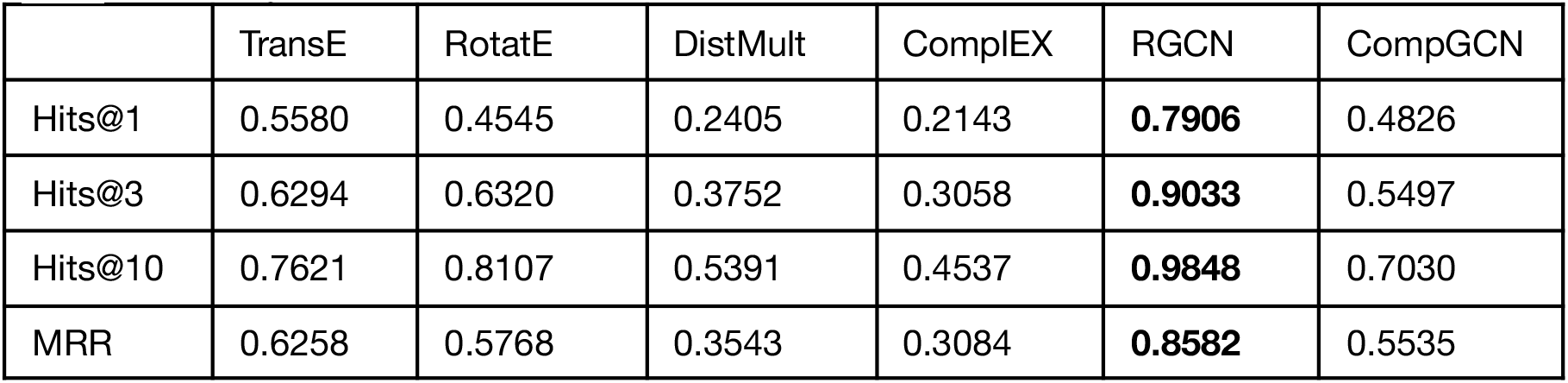
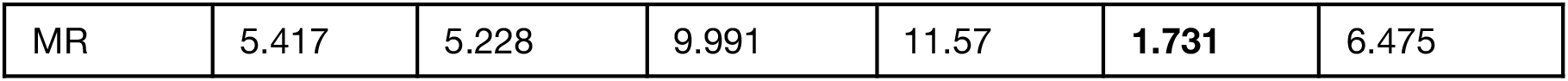
The metrics of link prediction results for different models on integrated knowledge graph, ADInt, by clinical trials dataset evaluation.

|  | TransE | RotatE | DistMult | Complex | RGCN | CompGCN |
| --- | --- | --- | --- | --- | --- | --- |
| Hits@1 | 0.5580 | 0.4545 | 0.2405 | 0.2143 | **0.7906** | 0.4826 |
| Hits@3 | 0.6294 | 0.6320 | 0.3752 | 0.3058 | **0.9033** | 0.5497 |
| Hits@10 | 0.7621 | 0.8107 | 0.5391 | 0.4537 | **0.9848** | 0.7030 |
| MRR | 0.6258 | 0.5768 | 0.3543 | 0.3084 | **0.8582** | 0.5535 |
| MR | 5.417 | 5.228 | 9.991 | 11.57 | **1.731** | 6.475 |

### Embedding representation of knowledge graph

Subsequently, we utilized t-SNE (t-distributed stochastic neighbor embedding) ^34^ to obtain two-dimensional projection of the learned node representations. t-SNE is a technique that reduces high-dimensional data to low-dimensional data while preserving the distribution properties of the original data. Moreover, it expresses the similarity between concepts through the proximity between nodes. As depicted in Figure 1, nodes with similar types tend to be grouped together, particularly the DS nodes.

**Figure 1:**
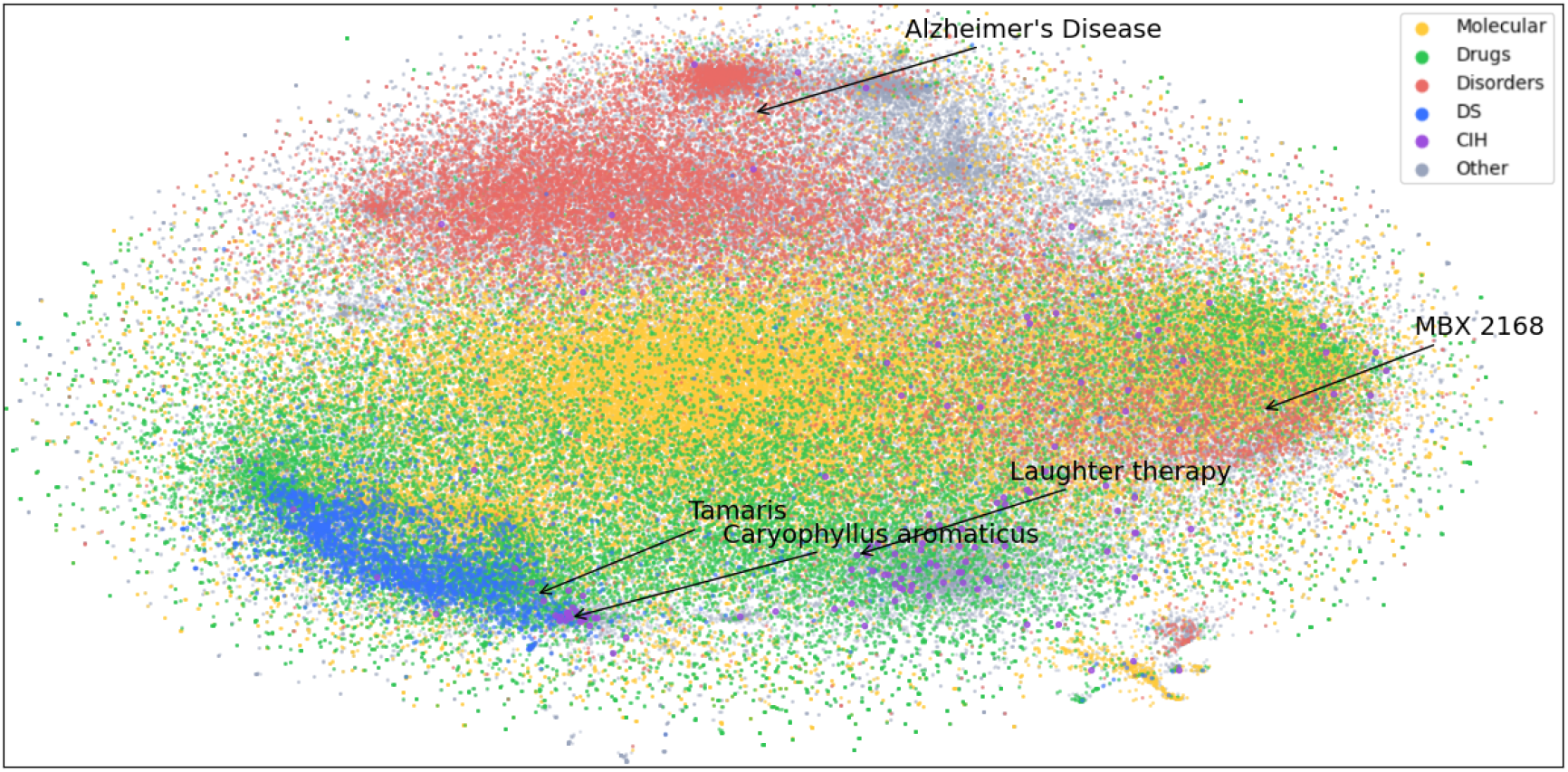
Visualization of nodes in ADInt dimensionally reduced by t-SNE algorithm and shown in a two-dimensional space. Different types of nodes are represented by different colors. Yellow: Molecular. Green: Drugs. Red: Disorders. Blue: DS (dietary supplement). Purple: CIH (complementary and integrative health). Gray: others.

### Discovered NPI list for AD prevention

We utilized the embedding information obtained from R-GCN to compute the score of each candidate triple. Specifically, we designated the tail node of these corrupted triples as C0002395 (Alzheimer’s Disease) and the edge as {PREVENTS, TREATS}. We then attempted to construct different triples by using all nodes in the graph as head nodes and calculated their score using the R-GCN model. Our focus was solely on the discovery of novel triples; thus, we excluded triples that already existed in ADInt. For novel triples, a higher score indicated a higher probability of being closely related to the true relationship. We categorized the triples into two groups based on the type of the head node, including DS and CIH, to discover novel NPIs for Alzheimer’s disease. The top 10 predicted novel candidates for Alzheimer’s disease are presented in Table 4

**Table 4:**
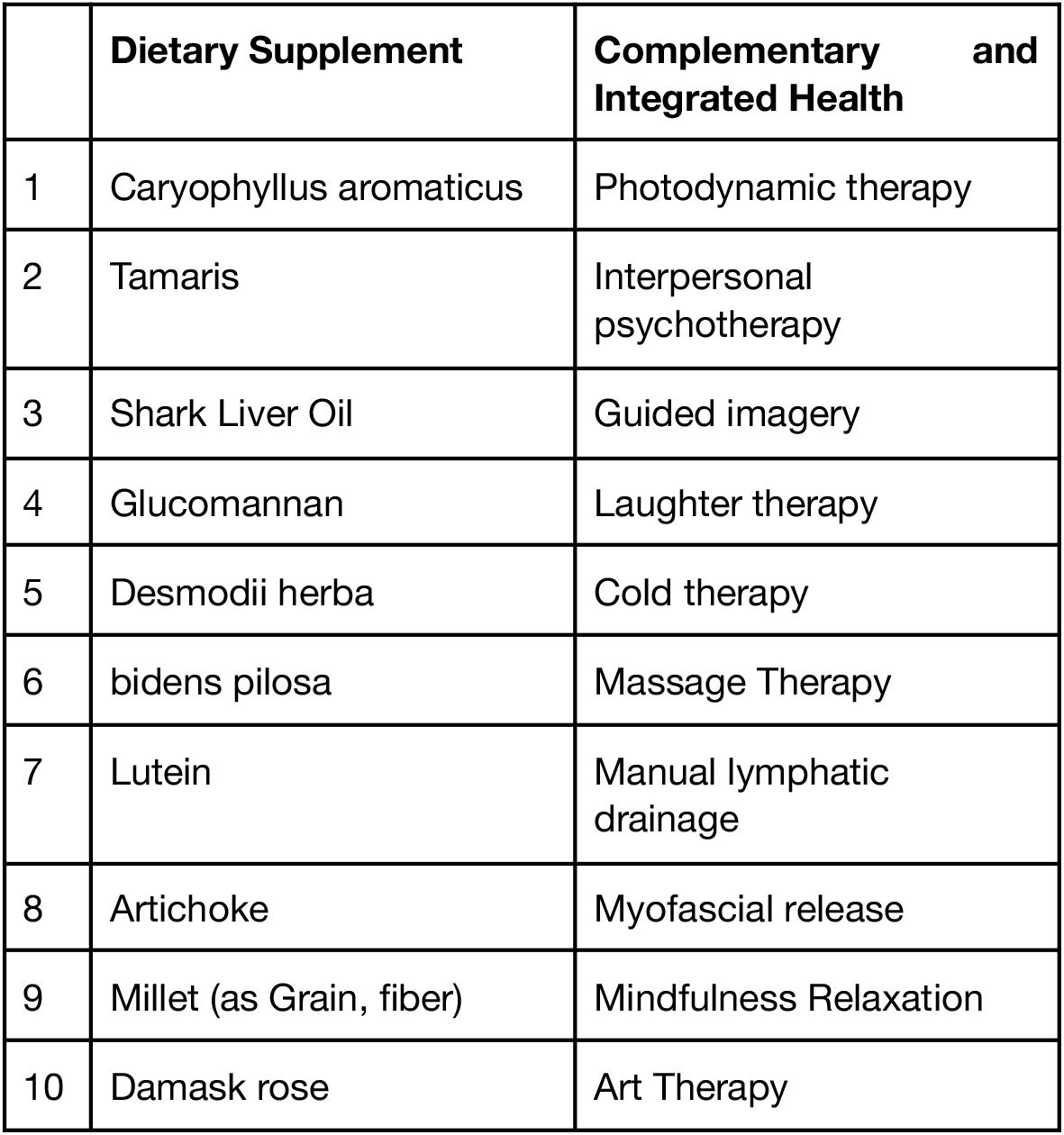
Top 10 proposed entities for different categories with predicate PREVENTS/TREATS.

|  | **Dietary Supplement** | **Complementary and Integrated Health** |
| --- | --- | --- |
| 1 | Caryophyllus aromaticus | Photodynamic therapy |
| 2 | Tamaris | Interpersonal psychotherapy |
| 3 | Shark Liver Oil | Guided imagery |
| 4 | Glucomannan | Laughter therapy |
| 5 | Desmodii herba | Cold therapy |
| 6 | bidens pilosa | Massage Therapy |
| 7 | Lutein | Manual lymphatic drainage |
| 8 | Artichoke | Myofascial release |
| 9 | Millet (as Grain, fiber) | Mindfulness Relaxation |
| 10 | Damask rose | Art Therapy |

Figure 2 displays the network structure of the top-ranked predicted results. The network highlights three pathways that include a set of interesting findings, which will be further discussed in the following sections. Specifically, this pathway reveals potential mechanisms through which CIH and DS may influence the risk of AD, and suggests potential targets for therapeutic interventions. The identified associations and pathways represent a promising direction for future research into the prevention and treatment of AD.

**Figure 2:**
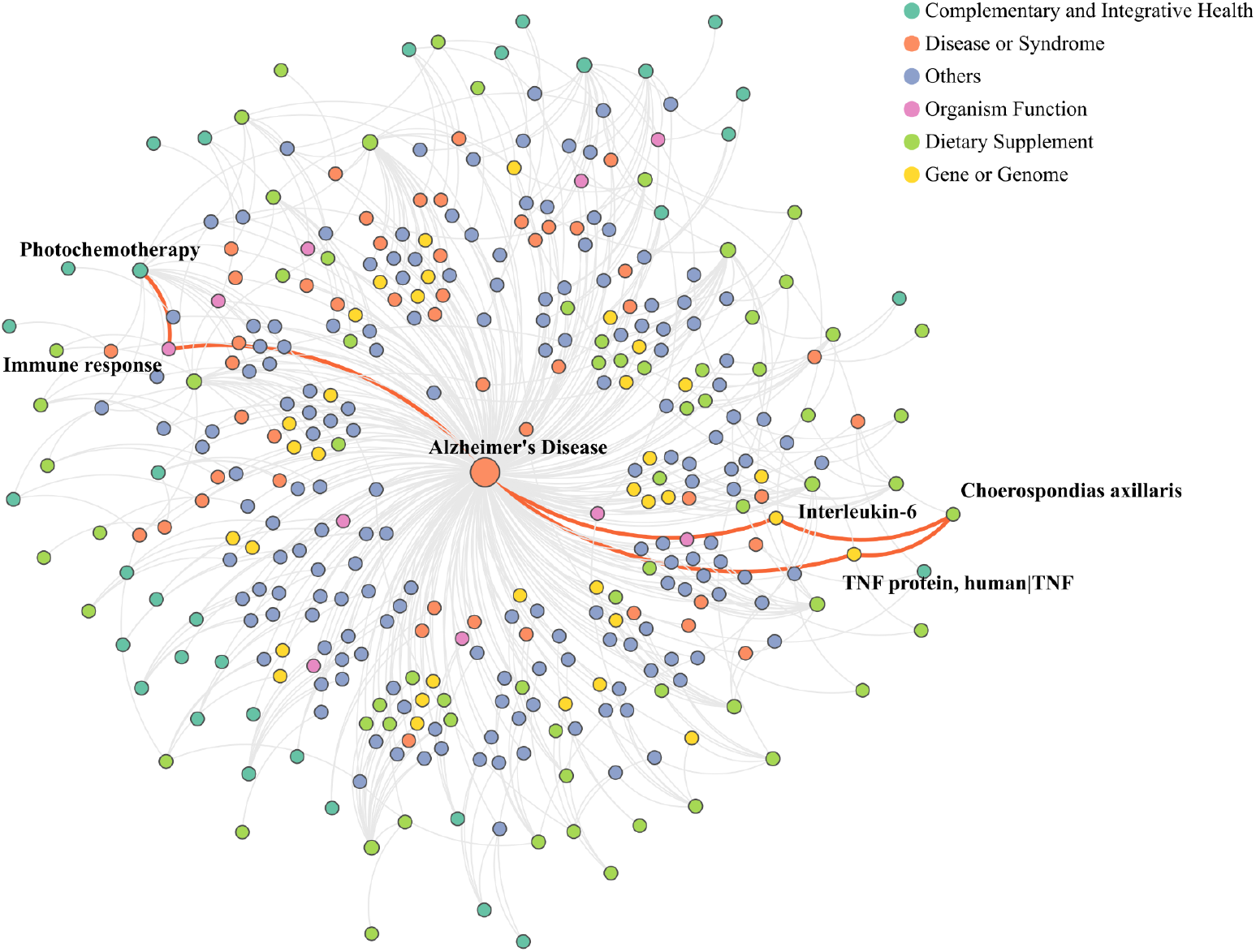
Top-ranked predicted results of ADInt-based exploration.

## Discussion

In this study, we compared various LP methods on the task of knowledge discovery. The R-GCN model has demonstrated superior performance over other models on both the Time Slicing and Clinical Trials test sets (see Table 2 and Table 3). Notably, TransE exhibited the second-best overall performance, which is consistent with our prior work ^30^ demonstrating that relatively simple TransE outperformed other knowledge graph embedding methods (RotatE, DistMult, ComplEX) on the extended SemMedDB. We speculate that the poor performance of DistMult and ComplEX is due to their preference for high-degree entities, which we removed during the data preprocessing stage ^35^. We believe that the reason for RotatE’s underperformance is similar, as our filtered knowledge graph emphasizes simple relations. Although RotatE addresses some of the limitations of TransE in handling multiple and symmetric relations by introducing complex spaces ^36^, our findings suggest that this approach may not be appropriate for our knowledge graph. The superior performance of R-GCN suggests that the neighborhood aggregation operation of the graph convolution network is useful for learning graph representations ^37^. However, we found that another graph convolutional network-based model, CompGCN, had a mediocre performance. We hypothesize that CompGCN’s reliance on linear transformations for relation embeddings does not suit our knowledge graph ^38^. Additionally, our evaluation of R-GCN on the Clinical Trials dataset, which mainly focuses on PREVENTS and TREATS relations, outperformed its performance on the Time Slicing evaluation. These results demonstrate that R-GCN is adept at distinguishing which subjects are feasible for treating or preventing AD. It is worth noting that while our experiments confirm R-GCN as the optimal LP model, metrics such as MR, MRR, and Hits@ratio only reflect the model’s ability to predict interventions being trialed or known interventions. Indeed, models with low metrics may still produce valuable results ^30^. Nevertheless, these metrics can inform model selection for NPI repurposing.

We used discovery patterns to generate mechanistic pathways for high-scoring triples predicted by the R-GCN model through the Neo4J platform. Photodynamic therapy (PDT) is a clinically used approach for treating various medical conditions, ranging from age-related macular degeneration to malignant tumors such as prostate cancer patients. PDT involves the use of light and a photosensitizing chemical substance along with molecular oxygen to elicit cell death ^39^. Recently, PDT has been proposed as a potential therapeutic option for AD ^39^. The precise mechanism of how PDT can provide therapeutic benefits for Alzheimer’s disease remains elusive, and the practical use of PDT for treatment of AD is basically non-existent given that tissue must be directly exposed to light, which is not feasible when dealing with the entire brain. However, this finding provides theoretical support for treating AD through modulation of the immune system. For instance, a study evaluating the use of PDT with 5-aminolevulinic acid on mice has reported that it affects the immune response ^40^. The study found that there was a significant reduction in the mRNA expression of interleukin-22 (IL-22), a cytokine produced by several immune cells that is associated with inflammation. Converging evidence has demonstrated that immune/inflammation response plays a crucial role in the initiation and regulation of Alzheimer’s disease ^41^. Thus, our PDT finding, while based on a therapy that has major practical limitations for treating AD, highlights immune mechanisms for preventing and treating AD. It should be noted that this is a preliminary finding based on a limited number of studies, and more research is needed to confirm these results.

Choerospondias axillaris, commonly known as Nepali hog plum, is a fruit that is approximately three centimeters long with sour flesh and yellow skin. Plums and other yellow-skinned fruits, such as papayas, tangerines, and oranges, are high in ß-cryptoxanthin, an antioxidant. A recent study ^42^ found an inverse association between serum β-cryptoxanthin levels and the incidence of Alzheimer’s Disease and all-cause dementias in individuals who consumed yellow-skinned fruits. Specifically, an increase of 8.6 micromole/liter in serum β-cryptoxanthin levels was associated with a 14% decreased risk of Alzheimer’s disease. To propose a potential mechanism for this protection, we examined the patterns between Choerospondias axillaris and Alzheimer’s disease. In a study ^43^, it was found that Choerospondias axillaris inhibits both TNF protein and interleukin-6. These two inflammation mediators are well-known inducers of Alzheimer’s disease, as demonstrated in previous studies ^44,45^. Specifically, interleukin-6 has been linked to the pathogenesis of Alzheimer’s disease, while tumor necrosis factor-α has been proposed as a potent therapeutic target for Alzheimer’s disease. Lutein, a carotenoid also found in Choerospondias axillas, we also found as a protective intervention. This finding corroborates prior reports that demonstrated an inverse association between lutein intake and dementia occurrence ^45^. Furthermore, increased lutein intake has been associated with lower levels of AD neuropathology postmortem ^46^. Overall, Choerospondias axillaris and other yellow-pigmented fruits may act as protectors by reducing the levels of pro-inflammatory cytokines crucially implicated in Alzheimer’s disease.

There are several possibilities for future improvements to our approach. Firstly, we augmented SuppKG with triples extracted from the SemMedDB database, indicating that all triples in our ADInt were obtained through literature-based discovery. In order to further enhance our knowledge graph, we can merge it with other comprehensive biomedical databases and biological networks, such as DrugBank and KEGG ^47^. This will enable us to expand the scope of our analysis and identify additional relevant interventions. Secondly, in addition to knowledge graph embedding and graph neural network models, other methods such as rule-based and reinforcement learning techniques have also demonstrated promising results on LP tasks. These methods could also be explored in future studies on drug repurposing. Lastly, since the determination of the plausibility of an intervention and its pathways to Alzheimer’s disease is a labor-intensive process, only the top 10 of each scoring table were evaluated by experts. However, in future work, larger samples could be considered if the necessary resources are available.

Our analysis emphasizes the growing importance and popularity of studying NPIs in the context of disease management. By demonstrating the efficacy of our approach in revealing intricate relationships between biomedical entities, particularly NPI entities, and diseases of interest, we provide plausible mechanistic explanations for these associations. Notably, our contributions in this field include creating valuable NPI resources and developing an innovative framework to predict NPIs that may potentially be repurposed for AD. To the best of our knowledge, this is the first study that specifically aims to discover NPIs for AD. Furthermore, the versatility and adaptability of our approach enable its application to NPI discovery for a wide range of other diseases, including COVID-19. Our proposed approach also holds significant potential in addressing various clinical questions, such as the discovery of drug adverse reactions and drug-drug interactions, further emphasizing the importance and applicability of our research in the broader biomedical field.

## Methods

The complete workflow is depicted in Figure 3. In order to investigate the association between PIs and NPIs and AD, we initially conducted preprocessing and integration of triples extracted from SemMedDB and SuppKG. Subsequently, we employed several graph representation models to derive the embedding information of ADInt, which included four knowledge graph embedding models (TransE ^33^, RotatE ^36^, DistMult ^48^ and ComplEX ^49^) and two graph convolutional network models (R-GCN ^50^ and CompGCN ^51^). Ultimately, we selected the most effective model for generating hypotheses regarding and NPIs for AD.

**Figure 3:**
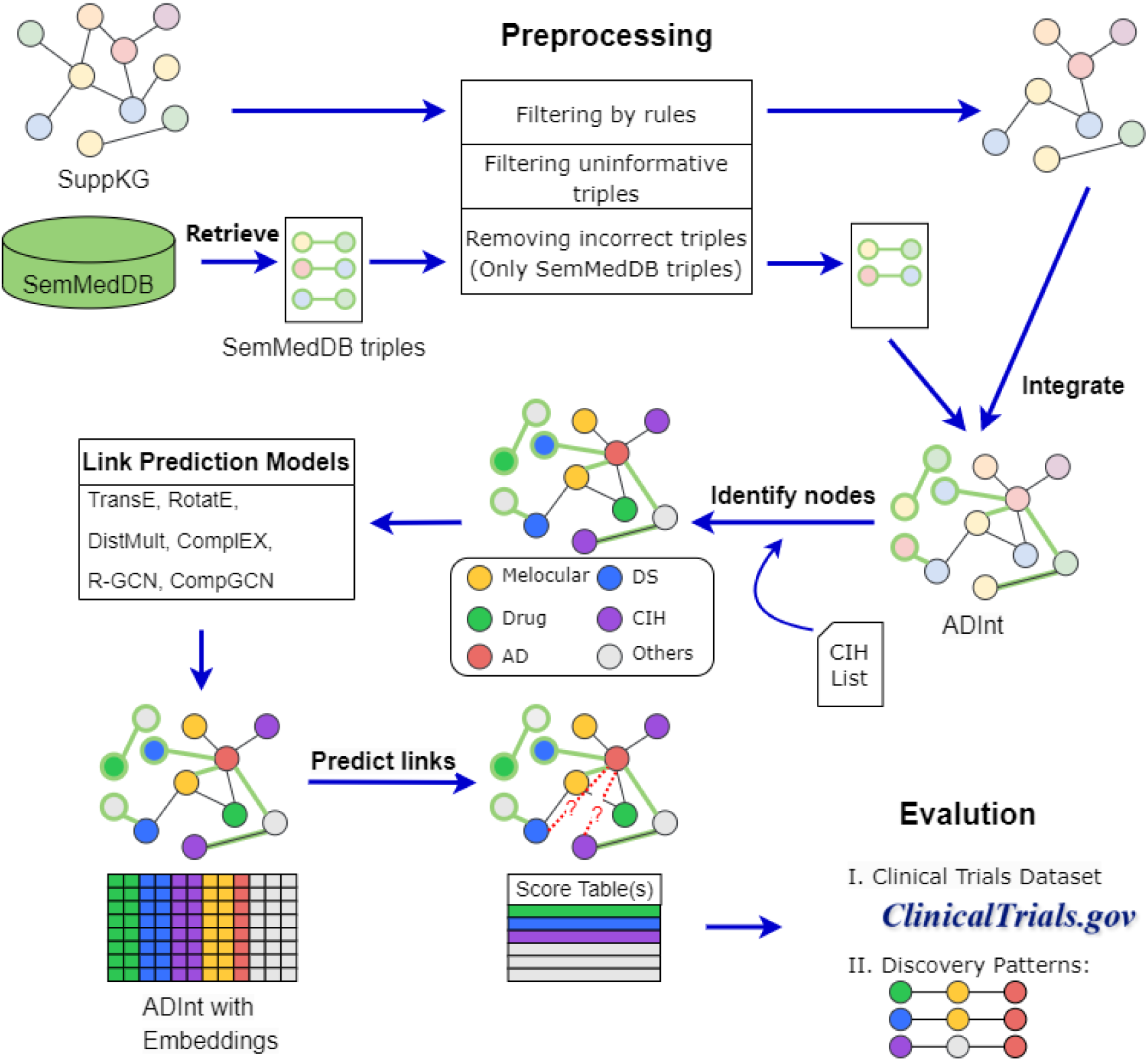
Diagram illustrating the workflow of the methodology.

### Materials

SuppKG ^27^ is a knowledge graph that focuses on DS and has been shown to be useful in discovering potential drug-supplement interactions (DSI) through discovery patterns. In this study, we utilized SuppKG to explore DS for AD. To create SuppKG, the domain of the conceptual space of MetaMap ^52^ used in SemRep ^53^ was extended by incorporating dietary supplement terminologies and relations contained in the Integrated Dietary Supplement Knowledge base (iDISK) ^54^. Subsequently, the extended SemRep was employed to extract semantic relations (triples) related to dietary supplements from PubMed abstracts retrieved using terms contained in iDISK. During the process of extracting semantic relations, some extracted semantic meanings were found to be opposite to the intended purpose of the corresponding text, resulting in the extraction of triples with opposite meanings. Due to the poor performance of SemRep (0.69 precision and 0.42 recall ^53^), a fine-tuned PubMedBERT model ^55^, which is a pre-trained Bert model with abstracts from PubMed, was utilized to eliminate incorrect triples. SuppKG comprises 56,635 nodes and 595,222 directed edges, including 2,928 DS-specific nodes and 164,738 edges. The nodes in SuppKG are identified by unique UMLS CUIs, while the predicates in UMLS Semantic Network label the edges. To easily distinguish the DS-specific nodes, a letter “D” was added before the CUI representing the concept of dietary supplements. For example, “DC0633482” was used to indicate that “myrtol” is a dietary supplement concept.

SuppKG is a valuable resource for discovering potential drug-supplement interactions through pattern discovery, but due to its focus on the dietary supplement domain and the limitations of its source data, it may not include all pathway information related to AD or other CIH approaches beyond dietary supplements. To address this limitation, we extended SuppKG with additional triples extracted from the SemMedDB database ^29^. SemMedDB is a repository of semantic triples extracted from PubMed abstracts and titles using the SemRep program ^53,56^, which provides detailed information such as the source text and PMID of the source article. We obtained triples from the PREDICTION table of SemMedDB and filtered them based on the correctness of the source sentences and text, which we extracted from the SENTENCE and PREDICATION_AUX tables. This allowed us to supplement SuppKG with a broader range of information related to interventions for AD beyond the dietary supplement domain.

### Preprocessing and Integration

In this study, an enhanced representation of nodes and relations in the knowledge graph is proposed by filtering out low-quality triples. Low-quality triples often describe generic facts that are not meaningful for the study. For instance, the triple (disease, AFFECTS, patient) with “disease” and “patient” being generic concepts is not useful in this study. Additionally, since the triples in SuppKG and entries in SemMedDB database are extracted from text using the SemRep text mining tool, some of the semantic relations expressed by the triples may not align with the intended meaning of the source text. Therefore, preprocessing is necessary to integrate the information from SuppKG and SemMedDB. The preprocessing steps can be classified into three categories ^30^:

#### 1) Filtering triples by rules

First, we removed nodes in the graph that represented generic concepts, which was done by referencing the GENERIC_CONCEPT table provided by the SemMedDB database. This table contained concepts such as “Disease” and “Cells,” which are known to be too broad to be useful for knowledge discovery. Additionally, concepts with semantic groups that were not likely to be useful for predicting interventions for ADRD were eliminated, including “Activities & Behaviors,” “Concepts & Ideas,” “Objects,” “Occupations,” “Organizations,” and “Phenomena.” Finally, only edges that were deemed relevant for LP were kept, specifically those with predicate types of AFFECTS, ASSOCIATED WITH, AUGMENTS, CAUSES, COEXISTS WITH, COMPLICATES, DISRUPTS, INHIBITS, INTERACTS WITH, MANIFESTATION OF, PREDISPOSES, PREVENTS, PRODUCES, STIMULATES, and TREATS.

#### 2) Removing high-degree concepts and uninformative semantic relations

High-degree High-degree concepts in the KG may be too general to be useful for knowledge discovery due to their broad associations with many other concepts. To address this issue, we first computed the out-degree 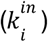 and in-degree 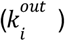 of each node in the KG. Next, we calculated a log likelihood measure known as *G*^2^ ^57^ for each triple, which quantifies the strength of the relationship between the items in the triple. The *G*^2^ formula is given by:

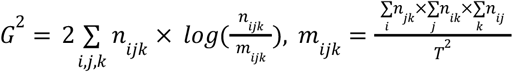

where *n* _*ijk*_ is the item *i,j,k* in the observation table (OT) containing observed frequencies of a triple, *m*_*ijk*_ is the item *i,j,k* in the expectation table (ET) describing the expected values assuming independence of terms in triples, and *T* = Σ *n*_*ijk*_. Finally, we normalized 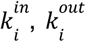 and *G*^2^ and summed them up together to get a final score for each triple. A higher score indicates that the triple is less specific and informative. For instance, the triple (Pharmaceutical Preparations, AFFECTS, Sleep) has a higher score and is more general compared to (DZIP1 gene, AFFECTS, heart valve development). Consequently, we filtered out some triples with high scores to manage the size of the knowledge graph for computational efficiency.

#### 3) Further removing incorrect triples by a trained PubMedBert model

The triples extracted from the SemMedDB database through SemRep may contain false positives, as the semantics expressed by the triples may differ or be contrary to the content of their source sentences. To address this issue, we utilized a PubMedBert binary classification model that was fine-tuned in our previous work to evaluate the correctness of the triples by referencing their source sentence ^30^. The F1 score of this model was 0.854, with a recall of 0.895 and a precision of 0.816.

For both SemMedDB and SuppKG triples, we applied steps 1) and 2) described above, but only applied step 3) to SemMedDB triples, as similar processing had been done during the generation of SuppKG. After filtering, we integrated the resulting triples from both sources, with DS concept nodes in SemMedDB triples identified by adding the letter D before their CUIs to match the identifiers in SuppKG. As the subject and object entities of the integrated triples are identified by UMLS CUIs and their predicates come from the UMLS Semantic Network, we added new triples to SuppKG that did not overlap with its existing triples, without mapping concepts or integrating ontologies. The resulting integrated knowledge graph, named ADInt, was obtained.

### NPI nodes identification

We employed multiple approaches to identify nodes representing drugs, DS, and CIH concepts in our knowledge graph for analysis and repurposing efforts for AD. For drug nodes, we can directly utilize the semantic types provided in the UMLS Metathesaurus. Specifically, we identify a node as a drug node if its semantic type is Pharmacologic Substance (phsu) or Organic Chemical (orch). However, identifying DS concept nodes based on their semantic type is not feasible. Nonetheless, in SuppKG, DS concept nodes are denoted by a special mark, a letter D added before their CUI. This mark was retained during the integration of SuppKG and SemMedDB triples, allowing us to easily identify these nodes as DS concepts. Unlike drug and DS nodes, nodes describing CIH concepts cannot be identified directly from the knowledge graph. To overcome this limitation, we utilized an external list of CIH concepts provided by a graduate student of informatics with a background in Medicine. This list, known as the CAM concepts list or CIHLex, was compiled based on a review of the literature, as described in our previous work ^58^ and Natural Medicines ^59^.

Given that DS and CIH nodes may have semantic types of phsu or orch, which are also associated with drug concepts, it is possible for overlap to occur. To address this issue, we prioritize the identification of DS or CIH concepts over drug concepts. If a node has been identified as either DS or CIH, it is considered as such, regardless of its semantic type of phsu or orch. This approach ensures that there is no ambiguity in the identification of nodes within the knowledge graph.

### Link prediction models training

A knowledge graph can be represented as a labeled directed multi-graph *KG* = (*E, R, G*), where *E* denotes the set of nodes representing entities, *R* denotes the set of edges representing relations, and *G* ⊆ *E* × *R* × *E* is a set of triples⟨*h, r, t*⟩, where h represents the head entity, *r* represents the relation, and *t* represents the tail entity. Link prediction (LP) is an essential task in knowledge graph completion, which aims to infer missing facts or relationships from the existing ones. Despite the vast amounts of information contained in knowledge graphs, they are often incomplete due to various factors, such as noise, missing data, and sparsity. Thus, LP methods seek to infer new triples that may not be explicitly represented in the knowledge graph, but which can be logically deduced from the existing ones. The objective of LP aims to predict the most probable entity or relation that completes (*h, r, ?*) (tail prediction), (*h, ?, t*) (edge prediction), or (*?, r, t*) (head prediction). Although new triples (*h’, r’, t’*) that describe additional facts may also exist in our knowledge graph, they are not present for some reason. LP for knowledge graphs can be represented as a ranking task, which aims to learn a prediction function that assigns higher scores to true triples and lower scores to false triples. To perform LP on our knowledge graph, we explored four knowledge graph embedding models (TransE, Rotate, DisMult and ComplEX) and two graph convolutional network models (R-GCN and CompGCN).

TransE ^33^ is a simple and effective model for LP, particularly for modeling one-to-one relations. In TransE, a triple (h, r, t) is represented as a translation from the embedding of the head entity h to the embedding of the tail entity t, with the relation r acting as the translation vector in the embedding space. This formulation implies that if a triple (h, r, t) exists, the embedding of entity h plus the representation of relation r should be close to the embedding of entity t. The TransE score function measures the plausibility of a triple and is defined as follows

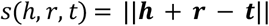

Where ***h, r, t*** ∈ ℝ^*d*^ is the embedding of *h, r* and *t*. Unlike TransE, The RotatE ^36^ model converts each relation to a rotation from a head entity to a tail entity in a complex vector space and the score function can be defined as

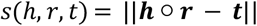

where ○ is a Hadamard product.

DistMult ^48^ is the most basic semantic matching models, and its scoring function can be defined as

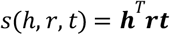

The drawback of DistMult is that it only works on symmetric relations, that is, the scores of (*h,r,t*) and (*t,r,h*) calculated by DistMult are the same. It may cause problems in our knowledge graph, for example the triple (Bariatric Surgery, TREATS, Alzheimer’s) and the triple (Alzheimer’s, TREATS, Bariatric Surgery) should have inconsistent scores. To address this limitation, ComplEX has been proposed as an extension of DistMult ^49^. ComplEX uses a complex vector space and is capable of modeling asymmetric relations. Specifically, head and tail embeddings of the same entity are represented as complex conjugates, which enables (*h, r, t*) and (*t, r, h*) to be distinguished. This allows ComplEX to provide consistent scores for both symmetric and asymmetric relations. The scoring function of ComplEX can be defined as follows

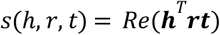

Where *Re*(·) is a real part of a complex vector.

GCNs are a neural network approach for processing graph-structured data ^60^. However, most existing GCNs are designed for simple undirected graphs and cannot handle the multiple types of nodes and directed links that exist in our knowledge graph. To address this challenge, we explored special graph convolutional neural network models that can handle heterogeneous graphs. Specifically, we evaluated two models: Relational Graph Convolutional Network (R-GCN) ^50^ and CompGCN ^51^. Based on the architectures of GCNs, R-GCNs ^50^ consider each different relation and perform feature fusion to participate in updating the hidden states of nodes. The propagation model for calculating the forward-pass update of a node in R-GCNs can be defined as

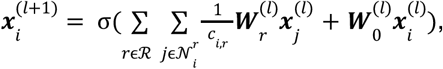

Where 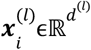 is the hidden state of *i*-th nodes in the *l*-th layer of the neural network; ℛ is the set of relations and 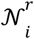 denotes the neighbor set of *i*-th node under relation *r*ϵℛ; 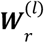 and 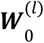 are the learnable weight matrix under relation *r* and self-loop weight matrix in the *l*-th layer respectively; *c*_*i,r*_ is a normalization constant that can either be learned or chosen in advance. Using R-GCNs for LP tasks can be regarded as a process of encoding and decoding: an R-GCN producing latent feature vectors of entities and a tensor factorization model exploiting these vectors to predict edges. Taking the DistMult decomposition as an example, the score of a triple (*h, r, t*) is calculated as ^50^

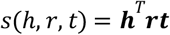

Thus, to make the model score observable triples higher than negative triples, the loss function can be defined as ^50^:

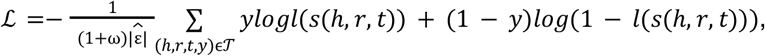

where 𝒯 is the set of all triples (including positive and negative triples); ω is the number of negative triples; 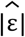 is the number of edges; *l*(.) is the logistic sigmoid function; and *y* is an indicator, where *y* = 1 means triple is positive, otherwise negative.

CompGCN ^51^ is another extended version of GCN for heterogeneous graphs, which systematically leverages entity-relation composition operations and jointly learning latent feature vector representations for both nodes and edges in the graph. Different from R-GCNs, CompGCN performs a composition operation Ф over each edge in the neighbor of central node through the embedding of edges and nodes. The update equation of nodes embedding in CompGCN can be defined as

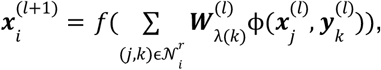

Where 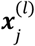 and 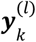 are the hidden state of neighboring *j*-th node and its *k*-th relation respectively in the *l*-th layer, and 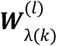 is a relation-type specific parameter, which can be used for direction specific weights. According to whether the edge is the original edge, inverse edge or self-loop edge, 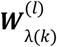 will correspond to different weight matrices. Ф(.) is used to aggregate two vectors of the same size, which can be Subtraction ^33^, Multiplication ^48^, or Circular-correlation ^61^. After updating the node embeddings, we can also update the relation embedding as follows ^51^

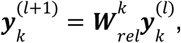

Where 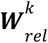 is a weight matrix that projects all relations to the same embedding space as nodes, which allows them to be used in the next layer. Similar to R-GCNs LP model, we select a tensor factorization model (convE) to calculate the score of triples. And the same standard binary cross entropy loss function is applied to training the convolutional networks.

All work was conducted using Python scripts. The implementation of the TransE, RotatE, DistMult, and ComplEX models was carried out with the DGL-KE 0.1.0.dev0 package ^62^ package. Both R-GCN and CompGCN models were constructed using the torch 1.13.1 ^63^ and DGL 1.0.1 ^64^ packages.

### Evaluations

#### Open LBD task

The open discovery approach is specifically aimed at generating innovative hypotheses; given a head node, the system produces associated tail nodes, thereby facilitating the identification of previously unexplored triple relationships. ^65^. In order to evaluate the effectiveness of our LP model, we utilized two evaluation methods. The first one is called Time Slicing ^66^. This evaluation approach involves partitioning the KG at a specific time and using the data prior to this time to train the model, and subsequently testing the model on the data following this time to determine if the links formed after the partitioning time can be accurately predicted. Specifically, in our work, we ordered the triples chronologically and divided the knowledge graph into training, validation, and testing sets in an 8:1:1 ratio, with earlier triples used for training and more recent ones for testing, where the date of publication of the paper mentioning the triple is used as its time, and the partitioning times were set as April 2020 and April 2021 respectively. To evaluate the model performance, we computed three metrics for each model: MR, MRR, and Hits@k (k = 1, 3, and 10). Specifically, for each true triple in the testing set, we generated a batch of negative samples by randomly replacing the head or tail nodes while ensuring that these negative samples do not exist in our graph, i.e., we employed corruption with filtering. We then used the trained model to calculate the scores for the true triple and its negative samples, and obtained the ranks of the true triples to obtain the metrics of MR, MRR, and Hits@k. MR represents the average rank assigned to the true relations in the test set, MRR is the average inverse rank of all true triples in the test set, and Hits@k is the percentage of relations in which the true triple appears in the top k ranked triples ^33^.

In the second evaluation approach, we utilized clinical trial data from ClinicalTrials.gov as a benchmark for predicting potential interventions for Alzheimer’s disease. Our approach was based on the assumption that interventions under investigation for AD have the potential to be repurposed for other indications. Specifically, we obtained a list of interventions utilized in AD clinical trials registered after April 21, 2020, by conducting a search for the term “Alzheimer” and restricting the results to interventional studies as of November 4, 2022. We excluded control interventions labeled as “placebo,” resulting in a total of 671 interventions. We processed these interventions using MetaMap with the UMLS 2022AA release to identify relevant UMLS concepts, resulting in 1606 concepts. These concepts were subsequently used as head nodes, with “TREATS” and “PREVENTS” serving as the relationships, and “Alzheimer’s disease” concepts as tail nodes, creating a series of new triples. Finally, we employed these newly generated triples based on clinical trial data as a test set to calculate MR, MRR, and Hits@k for each model.

#### Close LBD task evaluation

The closed discovery method strives to identify the connections between the given head and tail nodes in order to evaluate a specific hypothesis ^65^. Although the knowledge graph embedding and graph neural network models only provide node and edge representations, patterns from closed discovery were used to infer possible mechanisms for the repurposed interventions. To uncover potential logical connections between concepts in a network, we employed a closed discovery approach by combining sequences of relation types, such as “drug x INHIBITS substance y, substance y CAUSES disease z” ^32^. This method was used to identify possible pathways between nodes in the knowledge graph. For DS, The discovery patterns we focused on were:

~~~
**InterventionA**-INHIBITS|INTERACTS_WITH-**ConceptB** AND
**ConceptB**-AFFECTS|CAUSES|PREDISPOSES|ASSOCIATED-**Alzheimer’s disease** AND
NOT (**InterventionA**-TREATS|PREVENTS-**Alzheimer’s disease**)
~~~

where InterventionA is a node whose type is DS; ConceptB can be any concept; | indicates logical OR; and for Alzheimer’s disease, we focus on the node with identifier C0002395. To analyze the repurposing potential of Complementary and Integrative Health (CIH) interventions, we encountered a challenge due to the UMLS semantic types of most CIHs being “topp” (Therapeutic or Preventive Procedure) or “dora” (Daily or Recreational Activity). As these types do not have INHIBIT or INTERACT_WITH relationships to other concepts in the UMLS Semantic Network, and the number of possible paths is not extensive, we did not constrain the predicates in the patterns. The discovery patterns for CIH were:

~~~
**InterventionB**—(any predicate)-**ConceptB** AND
**ConceptB**-(any predicate)-**Alzheimer’s disease** AND
NOT (**InterventionB**-TREATS|PREVENTS-**Alzheimer’s disease**
~~~

where InterventionB is a node whose type is CIH. We visualized the network structure using ChiPlot (https://www.chiplot.online/).

## Data Availability

ADInt knowledge graph data is available in the following google drive: https://drive.google.com/drive/folders/187HnI2d-RRFeYk_C7MYSCHtVX-6IuNVS?usp=sharing. The complete SemMedDB database can be accessed directly on https://lhncbc.nlm.nih.gov/ii/tools/SemRep_SemMedDB_SKR.html.

https://drive.google.com/drive/folders/187HnI2d-RRFeYk_C7MYSCHtVX-6IuNVS?usp=sharing

https://lhncbc.nlm.nih.gov/ii/tools/SemRep_SemMedDB_SKR.html

## Code Availability

The code used for data preprocessing, model training, result evaluation and visualization in this study is available in the following repositories: https://github.com/YKXia0/LBD_AD.

## Acknowledgement

We would like to acknowledge the AWS Public Sector Cloud Credit for Research program to partially support this research.

